# Cardiac Abnormalities in Hispanic Women with Prior De Novo Hypertensive Disorders of Pregnancy

**DOI:** 10.1101/2023.08.24.23294595

**Authors:** Odayme Quesada, Shathiyah Kulandavelu, Catherine J. Vladutiu, Emily DeFranco, Margo B. Minissian, Nour Makarem, Natalie A. Bello, Melissa S. Wong, Maria Pabón, Alvin A. Chandra, Larissa Avilés-Santa, Carlos J. Rodríguez, C. Noel Bairey Merz, Tamar Sofer, Barry E Hurwitz, Gregory A. Talavera, Brian L. Claggett, Scott D. Solomon, Susan Cheng

**Affiliations:** Women’s Heart Center, The Christ Hospital Heart and Vascular Institute, Cincinnati, OH; The Carl and Edyth Lindner Center for Research and Education, The Christ Hospital, Cincinnati, OH; Department of Pediatrics, Miller School of Medicine, Miami, FL; Department of Obstetrics and Gynecology, University of North Carolina, Chapel Hill, NC; Division of Maternal Fetal Medicine, Department of Obstetrics and Gynecology, University of Cincinnati, Cincinnati, OH; Brawerman Nursing Institute, Cedars-Sinai Medical Center, Los Angeles, CA; Mialman School of Public Health, Columbia University Irving Center, NY, NY; Division of Maternal Fetal Medicine, Department of Obstetrics and Gynecology, Cedars-Sinai Medical Center, Los Angeles, CA; Division of Cardiovascular Medicine, Brigham and Women’s Hospital, Harvard Medical School, Boston, MA; University of Texas Southwestern Medical Center, Dallas, TX; National Institute on Minority Health and Health Disparities, Bethesda, MD; Albert Einstein College of Medicine, Bronx, NY; Barbra Streisand Women’s Heart Center; Department of Medicine, Brigham and Women’s Hospital, Harvard Medical School, Boston, MA; Department of Psychology, University of Miami, Miami, FL; South Bay Latino Research Center, Department of Psychology, San Diego State University, San Diego, CA

**Author notes:** **Address for Correspondence:** Odayme Quesada, MD, Women’s Heart Center, The Christ Hospital Heart, Vascular, and Lung Institute, Cincinnati, OH, Twitter: @OdaymeMD; and Susan Cheng, MD, MPH, Department of Cardiology, Smidt Heart Institute, Cedars Sinai Medical Center.

**Keywords:** Hypertensive disorders of pregnancy, women cardiovascular risk, left ventricular geometry, diastolic dysfunction, systolic dysfunction

## Abstract

**Background:** Hypertensive disorders of pregnancy (HDP) are associated with longer-term maternal risks for cardiovascular disease for reasons that remain incompletely understood.

**Methods:** The Hispanic Community Health Study/Study of Latinos (HCHS/SOL), a multi-center community-based cohort of Hispanic/Latino adults recruited 2008 to 2011 was used to evaluate the associations of history of de novo HDP (gestational hypertension, preeclampsia, eclampsia) with echocardiographic measures of left ventricular (LV) structure and function in Hispanic/Latina women with ≥1 prior pregnancy and the proportion of association mediated by current hypertension.

**Results:** The study cohort included 5,168 Hispanic/Latina women. Prior de novo HDP was reported by 724 (14%) of the women studied with an average age of 58.7 ± 9.7 years at time of echocardiogram and was associated with lower ejection fraction −0.66 (−1.21, −0.11), greater relative wall thickness (RWT) 0.09 (0, 0.18), and 1.39 (1.02, 1.89) greater odds of abnormal LV geometry after adjusting for blood pressure and other risk factors. The proportion of association mediated by current hypertension between HDP and LV ejection fraction was 0.09 (95% CI 0.03, 0.45), any abnormal LV geometry was 0.14 (0.12, 0.48), LV RWT 0.28 (0.16, 0.51), concentric LVH was 0.31 (0.19, 0.86), and abnormal LV diastolic dysfunction was 0.58 (0.26, 1.79).

**Conclusions:** In a large cohort of Hispanic/Latina women those with de novo HDP had detectable and measurable subclinical alterations in cardiac structure and both systolic and diastolic dysfunction that were only partially mediated by current hypertension.

## INTRODUCTION

The rates of hypertensive disorders of pregnancy (HDP), including preeclampsia and gestational hypertension, more than doubled from 2007 to 2019 in the United States (U.S.) with highest rates in Non-Hispanic Black and Hispanic/Latina women.^1,2^ Growing evidence demonstrates that history of HDP is associated with higher maternal risk for long-term cardiovascular disease (CVD) and CVD-related death; resulting in the addition of HPDs as risk-modifiers in the 2019 American College of Cardiology/ American Heart Association (ACC/AHA) primary prevention guidelines.^3–8^

Prior investigations have found evidence of structural cardiac abnormalities during the antepartum and immediate postpartum period, attributable in part to the short-term hemodynamic effects of pregnancy due to the stress of excess afterload.^9–12^ These cardiac structural changes, including increased left ventricular (LV) wall thickness, LV mass index, diastolic dysfunction and abnormalities in right ventricular (RV) strain, have been shown to persist postpartum.^11,13,14^ However, the role of chronic hypertension on these changes remains debatable.

Up to 20% of women with pregnancies complicated by HDP remain hypertensive after 6 months postpartum and have a 4-fold lifetime risk of chronic hypertension.^15–17^ Some studies show that adverse remodeling is driven by chronic hypertension regardless of HDP-history,^11,13^ whereas others demonstrate adverse remodeling associated with HDP is independent of development of chronic hypertension and a cumulative effect in those with both history of HDP and subsequent chronic hypertension.^14^ However, very little is known about the effects of HDP on cardiac abnormalities and the role of chronic hypertension in Hispanic/Latina women, one of the fastest-growing ethnic minority group in the U.S. with a diverse and broadly representative genetic architecture.^18^

The degree to which any important cardiac abnormalities occur well beyond the early postpartum period prior to the development of CVD decades later– notwithstanding the effects of postpartum or age-related chronic hypertension – has remained unclear. We hypothesize that history of de novo HDP is associated with pathologic alterations in cardiac structure and function that are only partially mediated by current hypertension. We aimed to examine this hypothesis in a diverse cohort of Hispanic/Latina women in the U.S.

## METHODS

### Study Sample

We studied participants of the Hispanic Community Health Study/Study of Latinos (HCHS/SOL), a multi-center community-based cohort of all Hispanic/Latino adults representing diverse backgrounds (Central American, Cuban, Dominican, Mexican, Puerto Rican, and South American).^18^ The HCHS/SOL sampling methods and design have been detailed previously.^19,20^ In brief, self-identified Hispanic/Latino men and women were recruited between March 2008 to June 2011 from 4 communities in the United States (Bronx, NY; Chicago, IL; Miami, FL; and San Diego, CA) using a multi-stage area probability sample design. At each stage of sampling, oversampling occurred, and sampling weights were generated to reflect the probabilities of selection. The institutional review board at each study site approved all protocols, and all study participants provided written informed consent. Data from the HCHS/SOL cohort is publicly available to researchers upon application to NHLBI BIOLINCC.

Of the 16,415 HCHS/SOL participants who enrolled in this study, we included women age ≥45 years who completed visit 2 and transthoracic echocardiography (TTE) and reported at least 1 prior pregnancy at baseline visit (2008-2011) or visit 2 (2014-2017). We excluded men (n=4,281), participants that did not complete visit 2 (n=4,792), did not complete echocardiogram (n=1,663), participants that were never pregnant or missing data on pregnancy history (n=405), and participants with missing data on HDP history (n=109). The final sample for this analysis included 5,168 women (724 with HDP and 4444 without HDP) (**Figure 1**).

**Figure 1.**
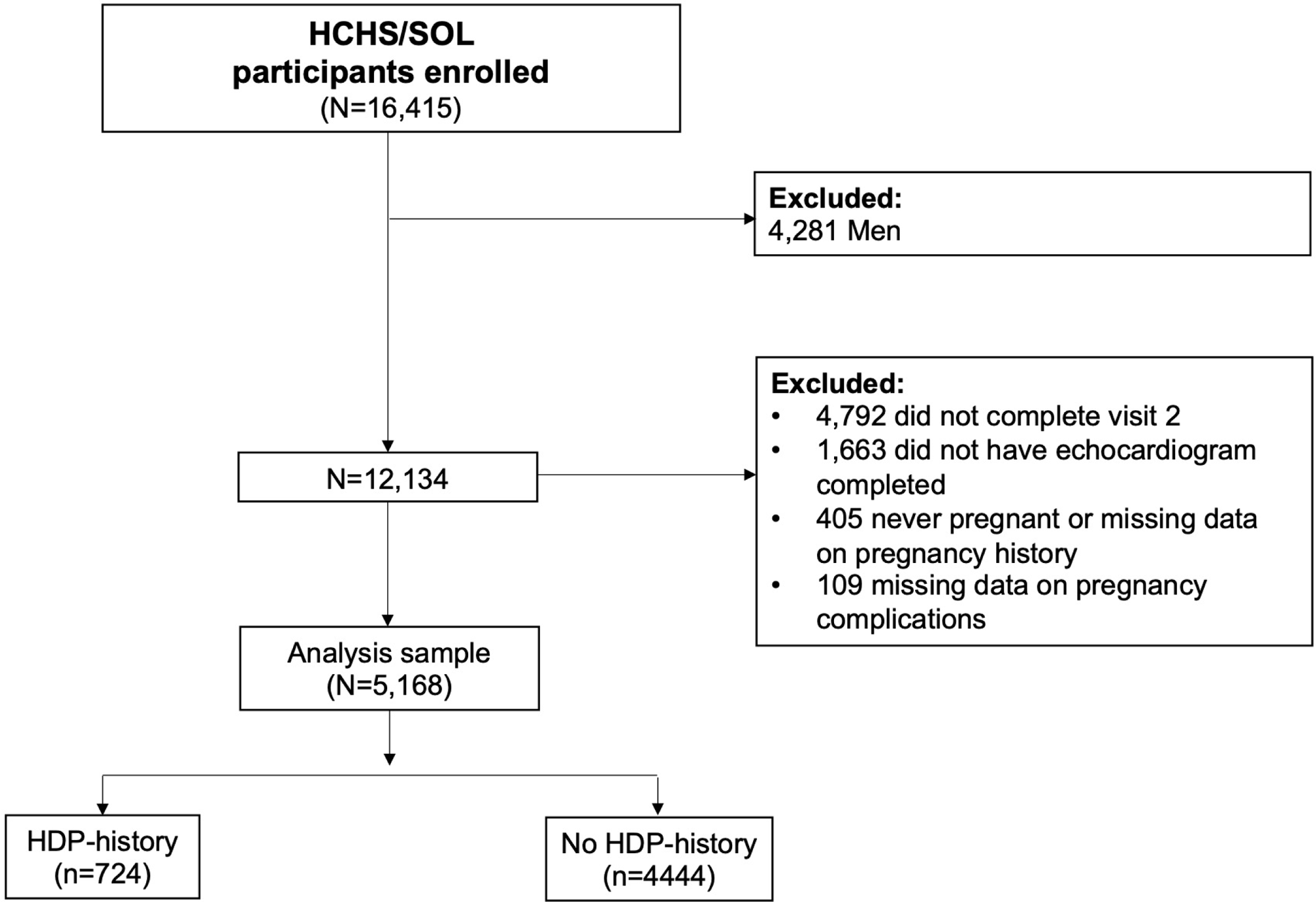
Sampling Strategy and Study Design. **Central Illustration.** Proportion of association between hypertensive disorders of pregnancy and measures of left ventricular structure and function mediated by current hypertension. Figure created using BioRender.

### Clinical and Echocardiographic Data Collection

At each study visit, all study participants underwent a standardized assessment of demographic and clinical characteristics including questionnaires regarding medical and pregnancy history along with standardized measurements of blood pressure (BP), as previously described.^20–22^ All participants included had self-reported data on the history of gestational hypertension, preeclampsia and eclampsia for all their pregnancies collected through questionnaires at visit 1 and 2. We defined composite HDP status as any history of de novo gestational hypertension, preeclampsia, or eclampsia.

At visit 2, participants age ≥45 years underwent comprehensive 2D TTE performed according to a previously detailed standardized protocol.^23^ In brief, TTE examination was performed with the participant in the partial left decubitus position with image acquisition techniques and measurements of cardiac structure and function performed according to American Society of Echocardiography guidelines.^24,25^ All image acquisition was performed by centrally trained and certified research sonographers and all imaging measurements were conducted by imaging technical specialists at the core HCHS/SOL Echocardiography Reading Center at Brigham and Women’s Hospital.^23^

As previously described, we defined concentric remodeling as LV mass index ≤95 gm/m^2^ and RWT >0.42, concentric LVH as LV mass index >95 gm/m^2^ and RWT >0.42, and eccentric LVH as LV mass index >95 gm/m^2^ and RWT ≤0.42.^24^ Any abnormal LV geometry was defined as presence of concentric remodeling, concentric LVH or eccentric LVH. Diastolic function was graded according to an algorithm that integrates American Society of Echocardiography (ASE) guidelines^26^ with Redfield criteria^27^ as previously described.^21,28^ In the analysis, LV diastolic dysfunction was dichotomized and grade I-IV diastolic dysfunction was compared to normal diastolic function.

### Statistical Analyses

We compared demographic, clinical, and echocardiographic traits in women with and without prior HDP using the Student’s t-test for continuous variables and Chi-square test for categorical variables. Holm-Bonferroni method was used to adjust for multiple comparisons. We then used multivariable-adjusted regression models to examine the association of prior HDP status with established measures of LV structure and function: LV ejection fraction, LV stroke volume, LV mass index, LV end-diastolic diameter, LV mass/end-diastolic volume ratio, LV relative wall thickness (RWT), peak tricuspid regurgitation velocity, lateral E/e’ ratio, abnormal LV geometry and LV diastolic function. We constructed linear and logistic regression models for continuous and categorical variables, respectively. For all echocardiographic traits, model 1 adjusted for age, study field center, and Hispanic/Latino background. Model 2 adjusted for the covariates in model 1, in addition to SBP and treatment with antihypertensive therapy at the time of TTE (Visit 2). Model 3 adjusted for the covariates in model 2, in addition to body mass index, diabetes, smoking, total number of prior pregnancies, total cholesterol/HDL ratio, and urine albumin-to-creatinine ratio all assessed at the time of TTE. Covariates were selected based on prior studies demonstrating association with HDP and/or LV measures of structure/function.^11,13,14^ No adjustments were made for multiple testing. Stratified analyses were performed by type of HDP for gestational hypertension and preeclampsia, however the sample size was too small to perform adjusted models for eclampsia. Reported values were survey-weighted, to account for the complex study design and the non-responses for visit 2.^19^ Weights were trimmed and calibrated to the 2010 Census characteristics by age, sex, and Hispanic/Latino background. In secondary analyses, we examined the extent to which current hypertension mediated the associations of HDP with echocardiography traits. Models are adjusted for age, field center, Hispanic/Latin background, and current hypertension stage 2 (defined as BP >140/90 mmHg or antihypertensive therapy). These analyses test the extent to which current hypertension mediate the associations of HDP with the given echocardiographic trait, whereby a mediation effect of 0 would indicate that current hypertension does not mediate the association and a mediation effect of 1 would indicate that current hypertension mediates all of the association (range of possible effect is from 0 to 1). We also assessed for potential interaction of current hypertension on HDP associations with each of the echocardiographic traits.

We considered statistical significance as a two-tailed P value and Holm-Bonferroni adjusted P value of less than 0.05. All statistical analysis were conducted using R (v4.0.4).

## RESULTS

Of the total study sample including 5,168 women with prior pregnancy, 724 (14%) reported a history of de novo HDP including 439 (61%) gestational hypertension, 219 (30%) preeclampsia, and 66 (9%) eclampsia. The mean age at the time of examination and TTE was 58.7±9.7 years. The demographic and clinical characteristics of the study sample at visit 2 are shown in **Table 1**. Women with a history of HDP, compared to those without HDP were younger, had a higher level of education, greater body mass index, and more frequent dyslipidemia. After the Holm-Bonferroni adjustment, dyslipidemia and HDL-cholesterol levels were no longer statistically different between the groups. With respect to echocardiographic LV characteristics (**Table 2**), women with prior HDP had lower LV ejection fraction, higher LV stroke volume, greater LV mass index, greater LV RWT, and higher peak tricuspid regurgitation velocity (PTRV). Accordingly, HDP was also associated with greater concentric LVH, presence of any abnormal LV geometry, and presence of abnormal diastolic function. After the Holm-Bonferroni adjustment, LV mass index and PTRV were not significantly different between the groups.

**Table 1.**
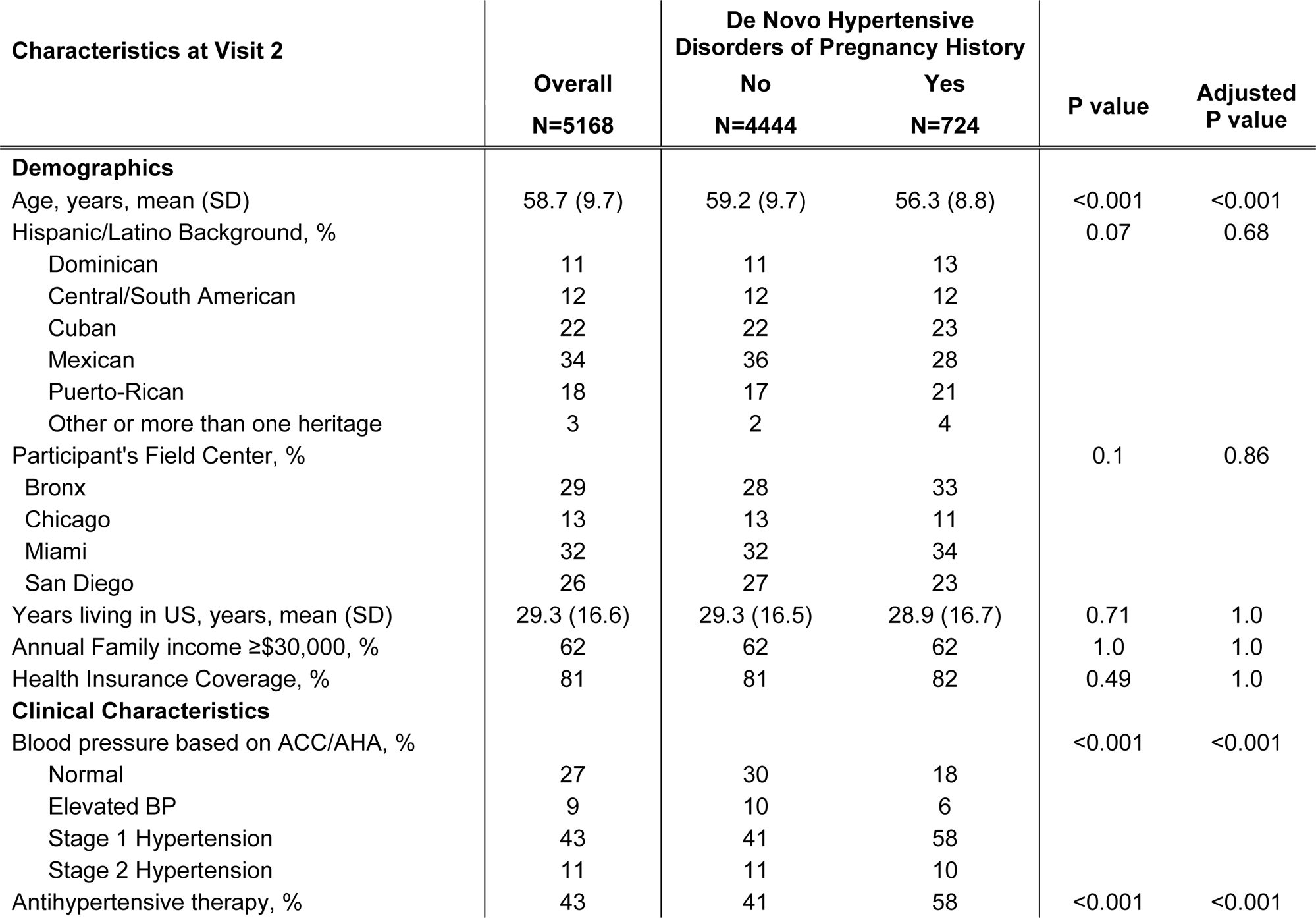
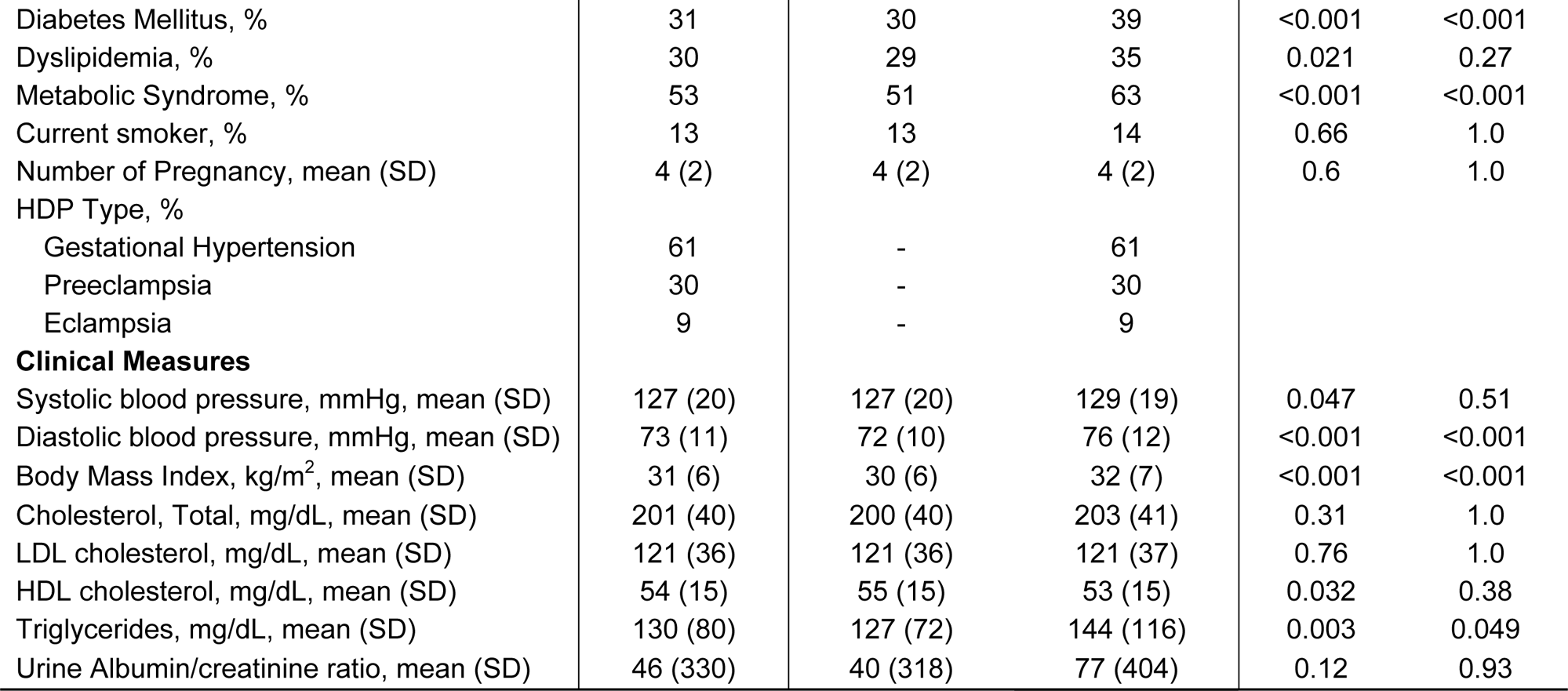
Demographics and Clinical Characteristics by De Novo Hypertensive Disorders of Pregnancy History.

**Table 2.**
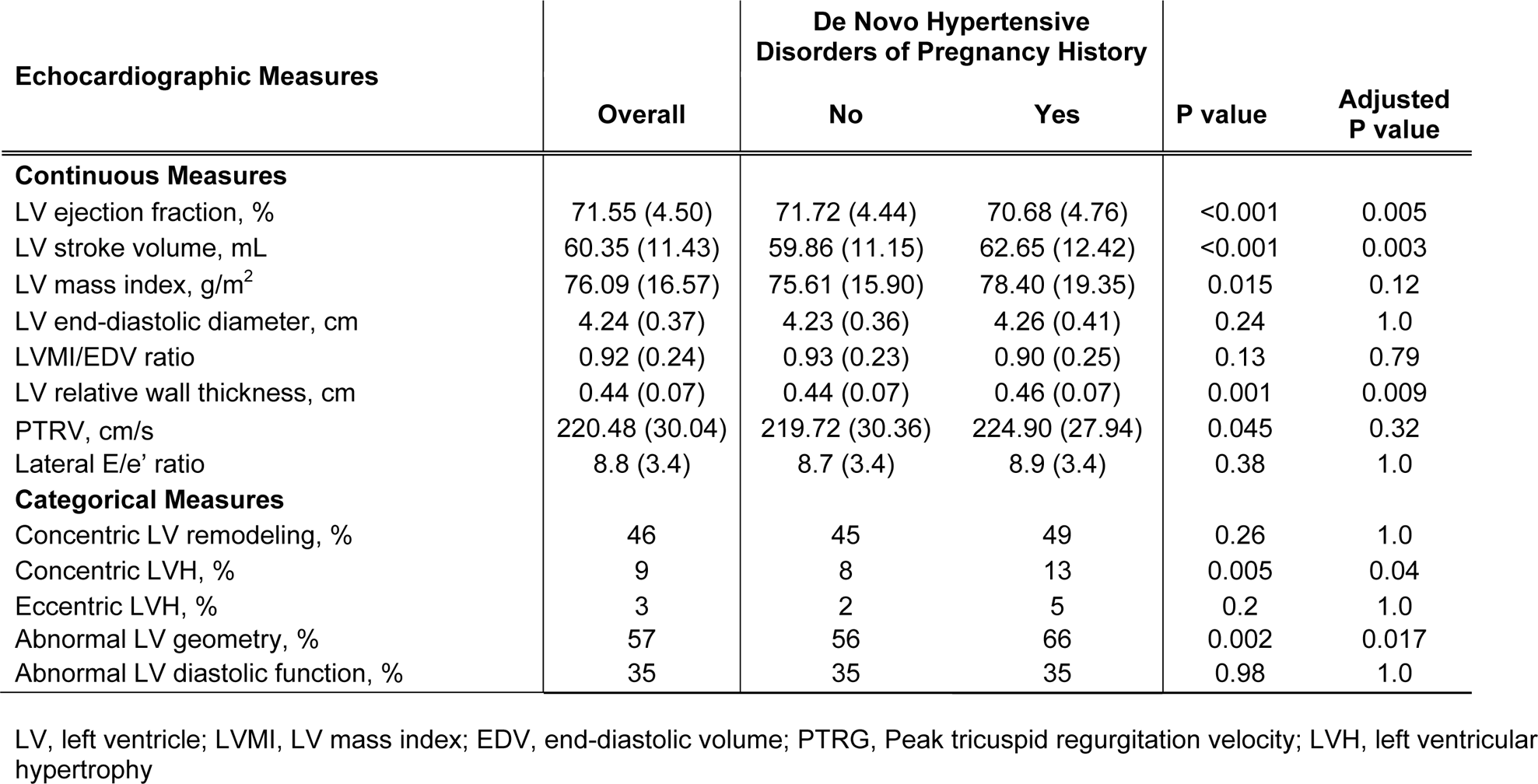
Differences in Measures of Left Ventricle Structure and Function by De Novo Hypertensive Disorders of Pregnancy History.

In multivariable-adjusted analyses of the continuous LV structure and function measures, prior HDP was associated with lower LV ejection fraction in all models including adjustment for systolic BP, antihypertensive therapy and traditional risk factors (P≤0.02) (**Table 3**). Prior HDP was also associated with increased LV RWT in all models (P≤0.039). Accordingly, among the categorical LV measures, prior HDP was associated with presence of concentric LVH in model 1 adjusted for age and demographic factors (P=0.001), but the association was attenuated in analyses adjusting for systolic BP and antihypertensive therapy (P=0.07). Notably, prior HDP was significantly associated with presence of any abnormal LV geometry in all models (P≤0.038). In addition, prior HDP was associated with presence of abnormal LV diastolic function in models adjusting for age and demographic factors (P=0.006); this association was attenuated in analyses adjusting for systolic BP and antihypertensive therapy (P=0.43). All other measures of LV structure and function were nonsignificant.

**Table 3.** Associations between De Novo Hypertensive Disorders of Pregnancy History and Measures of Left Ventricle Structure and Function.

| Echocardiographic Measures | Model 1 |  | Model 2 |  | Model 3 |  |
| --- | --- | --- | --- | --- | --- | --- |
|  | Coefficient (95% CI) | P value | Coefficient (95% CI) | P value | Coefficient (95% CI) | P value |
| **Continuous Measures** |  |  |  |  |  |  |
| LV ejection fraction, % | -0.90 (-1.45, -0.04) | 0.001 | -0.75 (-1.30, -0.21) | 0.007 | -0.66 (-1.21, -0.11) | 0.02 |
| LV stroke volume, mL | 1.72 (0.32, 3.12) | 0.016 | 0.94 (-0.41, 2.29) | 0.17 | 0.32 (-0.96, 1.61) | 0.62 |
| LV mass index, g/m<sup>2</sup> | 3.70 (1.46, 5.94) | 0.001 | 1.57 (-0.50, 3.64) | 0.14 | 1.05 (-1.01, 3.10) | 0.32 |
| LV end-diastolic diameter, cm | 0.01 (-0.49, 0.52) | 0.96 | -0.12 (-0.62, 0.38) | 0.65 | -0.29 (-0.75, 0.18) | 0.23 |
| LVMI/EDV ratio | 0.01 (-0.03, 0.04) | 0.72 | -0.01 (-0.04, 0.03) | 0.72 | 0 (-0.04, 0.03) | 0.92 |
| LV relative wall thickness, cm | 0.22 (0.13, 0.31) | <0.001 | 0.14 (0.05, 0.23) | 0.003 | 0.09 (0.00, 0.18) | 0.039 |
| PTRV, cm/s | 7.37 (2.50, 12.25) | 0.003 | 4.0 (-0.92, 8.92) | 0.11 | 2.22 (-2.65, 7.09) | 0.37 |
| Lateral E/e' ratio | 0.58 (0.19, 0.96) | 0.004 | 0.19 (-0.19, 0.58) | 0.33 | 0 (-0.37, 0.38) | 0.10 |
| **Categorical Measures** |  |  |  |  |  |  |
| Concentric LV remodeling, % | 1.33 (1.01, 1.75) | 0.046 | 1.25 (0.94, 1.67) | 0.12 | 1.16 (0.87, 1.55) | 0.31 |
| Concentric LVH, % | 1.92 (1.32, 2.79) | 0.001 | 1.45 (0.97, 2.16) | 0.07 | 1.30 (0.85, 1.99) | 0.22 |
| Eccentric LVH, % | 1.60 (0.80, 3.21) | 0.19 | 1.32 (0.64, 2.75) | 0.45 | 1.38 (0.63, 3.00) | 0.42 |
| Abnormal LV geometry, % | 1.86 (1.39, 2.49) | <0.001 | 1.51 (1.12, 2.03) | 0.007 | 1.39 (1.02, 1.89) | 0.038 |
| Abnormal LV diastolic function, % | 1.52 (1.13, 2.05) | 0.006 | 1.14 (0.83, 1.57) | 0.43 | 1.02 (0.73, 1.42) | 0.90 |

LV, left ventricle; LVMI, LV mass index; EDV, end-diastolic volume; PTRG, Peak tricuspid regurgitation velocity; LVH, left ventricular hypertrophy
Model 1 is adjusted for age, field center, Hispanic background; Model 2 is adjusted for all variables in Model 1, in addition to systolic blood pressure and antihypertensive therapy at Visit 2; Model 3 is adjusted for all variables in Model 2, in addition to diabetes status, smoking, total cholesterol HDL ratio, number of pregnancies, BMI, urine albumin-to-creatinine ratio

In secondary analyses, we examined the extent to which current hypertension mediated the associations of HDP with echocardiography traits. We found that the proportion of association between HDP and LV ejection fraction that was mediated by current hypertension was modest at 0.09 (95% CI 0.03, 0.45), with similar results seen for any abnormal LV geometry (0.14 [0.12, 0.48]). The proportions of association between HDP and other traits such as LV RWT (0.28 [0.16, 0.51]), concentric LVH (0.31 [0.19, 0.86]), and abnormal LV diastolic dysfunction (0.58 [0.26, 1.79]) that were mediated by postpartum hypertension were higher, as expected given results of multivariable-adjusted models described above (**Table 4**). We also assessed for potential interaction of current hypertension on HDP associations with each of the echocardiographic traits and found no significant interactions (P>0.30 for all). We observed no material difference in any results of analyzing data using survey-weighted versus unweighted values.

**Table 4.** Causal Mediation Analysis to Assess Mediation Effect of Current Hypertension for Observed Association Between De Novo Hypertensive Disorders of Pregnancy and Measures of Left Ventricle Structure and Function.

| **Echocardiographic Measures** | **Proportion of Association with HDP Mediated by Current Hypertension (95% CI)** | **P value** |
| --- | --- | --- |
| LV ejection fraction | 0.09 (0.03, 0.45) | 0.01 |
| LV relative wall thickness | 0.28 (0.16, 0.51) | <0.001 |
| Concentric LVH | 0.31 (0.19, 0.86) | <0.001 |
| Abnormal LV geometry | 0.14 (0.12, 0.48) | <0.001 |
| Abnormal LV diastolic function | 0.58 (0.26, 0.79) | 0.006 |

Models are adjusted for age, field center, Hispanic/Latino background, and current hypertension (defined as BP >140/90 or antihypertensive therapy).

In stratified analysis by type of HDP, there was a trend towards lowest ejection fraction and LV stroke volume and highest LV RWT in women with eclampsia history (**Supplemental Table 1**). The presence of concentric LVH was highest in women with history of gestational hypertension and presence of abnormal LV diastolic function was highest in those with history of preeclampsia and gestational hypertension. In multivariable-adjusted analyses, gestational hypertension was associated with LV RWT and 1.79-fold higher risk of abnormal LV geometry across all models, whereas there was no association between preeclampsia and any of the measures of LV structure and function (**Supplemental Table 2**).

## DISCUSSION

In our study of over 5,100 Hispanic/Latina women with prior pregnancy, 14% reported de novo HDP. Women with history of de novo HDP were significantly more likely to have measurable abnormalities in cardiac structure and function, including lower LV systolic function and higher rates of abnormal LV geometry than women without history of HDP. These cardiac alterations were in part mediated by the effects of current hypertension (**Central Illustration**).

The rate of de novo HDP in this study is consistent with the U.S. National Inpatient Sample between 2017 and 2019 which reported 12.5% (95% CI 12.2-12.8).^29^ To date, a scant number of studies have examined cardiac phenotypes up to a decade or longer following delivery prior to development of clinical CVD; these few studies, each involving sample sizes of less than 200 women with history of HDP, have found evidence of LVH or diastolic dysfunction.^30,31^ In studies accounting for chronic hypertension, findings have also been mixed on whether or not the changes in LV structure and function associated with HDP history are present after adjusting for chronic hypertension – an obvious potentially confounding contributor to subsequent cardiac risk.^11,13,14^ We extend previous work and analyze one of the largest samples of de novo HDP in 724 women and compared their cardiac phenotypes with 4444 similarly aged clinical controls (i.e. women with prior pregnancy but no HDP). Importantly, we found that prior HDP was associated with higher LV RWT and abnormal LV geometry – even after adjusting for current hypertension and CV risk factors – consistent with a recent study by Countouris et al.^14^ in a cohort of NHW and Black women. Our study extends these findings to Hispanic/Latina women who are underrepresented in research studies.

In this study history of pregnancy complicated by de novo HDP was associated with higher risk of abnormal LV geometry (defined as concentric LV remodeling, concentric LVH, or eccentric LVH as determined by LVMI and RWT) even after adjusting for important confounders. These findings are of clinical significance because abnormal LV geometry, particularly LVH, is an independent predictor of adverse CVD events including heart failure, ischemic heart disease, and sudden cardiac death.^32–34^ We also found an association between de novo HDP and abnormal LV diastolic function, which is linked with higher incidence of CVD events in healthy cohorts and is a strong predictor of progression to heart failure with preserved ejection fraction (HFpEF).^35,36^ Further, individual measures of LV geometry (LVMI, LV RWT) that we found to be associated with history of de novo HDP have also been shown to be associated with CVD events. For instance, in the Framingham Heart Study of Offspring cohort among adults free of CVD, each 10 g/m2 increment in LVMI was associated with 33% increased risk of CVD and each 0.1 unit in LV RWT was associated with 59% increased risk of CVD.^37^

Further, this is the first known study to identify evidence of HDP associated with decrements in LV systolic function. The finding of slightly lower LV ejection fraction despite greater concentric remodeling and a trend towards greater stroke volume suggests a type of contractile inefficiency that is not typically seen in the setting of current hypertension alone, suggesting a pathophysiology distinct form of hypertensive heart disease. These findings and the diastolic dysfunction exhibited by these mothers all suggest a pre-HFpEF remodeling phenotype, supported by the excess risk of HFpEF in women with preeclampsia, that warrants further investigation.^38,39^

Hypertension results in chronic central pressure overload and myocardial ischemia that leads to the development of LVH and heart failure. Therefore, we evaluated the extent to which current hypertension mediated the association between history of de novo HDP and measures of LV structure and function. Hypertension is associated with significantly higher rates of concentric LVH and is a strong independent predictor of LV diastolic dysfunction, but the association with eccentric LVH and LV systolic dysfunction is less robust.^40,41^ This helps explain why hypertension was a moderate mediator in the association between HDP and concentric LVH and LV diastolic dysfunction but a weaker mediator of LV ejection fraction (a measure of LV systolic function). Our results are consistent with large epidemiologic data indicating that the association of HDP with later-life CVD is only partially mediated by current hypertension.^5^ These findings provide insights into HDP as a potential novel mechanism to explain the disproportionally higher risk of heart failure in women with a history of HDP.^42,43^

The mechanisms by which HDP may lead to abnormalities in cardiac structure and function beyond the effects of current hypertension remain to be elucidated.^44^ Diabetes was found to be higher in women with history of de novo HDP and although we included diabetes in our adjusted models it may have contributed to some of our findings. In fact, glucose intolerance, insulin resistance, and diabetes have been shown to be associated with increased LV mass and wall thickness and reduced end-diastolic volume, stroke volume, and ejection fraction.^45–47^ Further, endothelial dysfunction is a hallmark of the pathophysiology underlying preeclampsia and some studies suggest this can persist after delivery.^48,49^ Activation of pathways involving the antiangiogenic soluble fms-like tyrosine kinase 1 (sFlt-1) in preeclampsia, which can remain at least modestly elevated postpartum, can contribute to endothelial dysfunction and deranged lipid metabolism.^50–52^ Dysregulations of the renin-angiotensin system may also contribute to persistent postpartum cardiac abnormalities. Additionally, shared upstream factors such as CV risk factors and/or genetics may predispose women to both HDPs and later in life pathological LV remodeling and function. Further investigations are needed to identify the potential mechanisms contributing to cardiac abnormalities and how these may be different by race and ethnicity.

### Study Limitations

Several limitations of our study merit consideration. The cross-sectional design of our study precludes inference of causality, although timing of reported prior HDP consistently preceded timing of assessed cardiac traits. Prior HDP status was based on self-report, which is subject to recall bias and limits precision with respect to subtypes of HDP; nonetheless, self-report has been evaluated as valid and thus applied in the vast majority of cohort studies on HDP.^53,54^ Data on severity of preeclampsia, number of pregnancies complicated by HDP, time period between HDP pregnancy and echocardiography, and length of postpartum hypertension is not available. Additionally, multiple echocardiographic measures were analyzed, which can lead to heightened Type 1 error rate. Despite these limitations, our study offered several strengths including analyses from the largest study to date investigating the relations of prior HDP with cardiac traits in Hispanic/Latina women who represent a demographically important yet historically understudied population. In addition, all cardiac traits were assessed from echocardiographic protocols that involved standardized image acquisition and centralized measures performed at a core center with high inter- and intra-reader reproducibility and BP obtained in a standardized method.

### Conclusions

In summary, we found in a large cohort of previously pregnant Hispanic/Latina women that those with history of de novo HDP had detectable and measurable pathological alterations in cardiac structure and systolic and diastolic dysfunction. Our findings suggest that women with prior HDP have pathophysiologic cardiac sequelae decades later, that likely play a role in modulating long-term CV risk. Notwithstanding the need for further investigations into the mechanisms driving HDP pathophysiology, our findings highlight the potential importance of targeted surveillance and interventions aimed at preventing later-life CVD events in these at-risk understudied women.

### Perspectives

Hispanic/Latina women are disproportionally affected by HDP, yet very little is known on how HDP affects cardiac structure and function in this growing population in the U.S. In this study of a diverse cohort of Hispanic/Latina women, those with history of de novo HDP had higher rates of abnormal LV geometry, and alterations in LV geometry (LVMI, RWT) and function (LVEF, diastolic dysfunction), known predictors of CV events and mortality. Notably, these findings appear to be in part mediated by current hypertension – underscoring the importance of screening for and managing hypertension in this cohort. Hypertension alone did not account for all the associations between history of de novo HDP and morphologic and functional cardiac alterations. With the higher rates of heart failure, particularly HFpEF, in women and the alterations in cardiac structure and function identified in this study we hypothesize HDP may be a sex-specific risk factor that warrants further investigation.

## Data Availability

Data from the HCHS/SOL cohort is publicly available to researchers upon application to NHLBI BIOLINCC.

## ABBREVIATIONS

BP: blood pressure
CVD: Cardiovascular disease
HCHS/SOL: Hispanic Community Health Study/Study of Latinos
HDP: Hypertensive disorders of pregnancy
LV: Left ventricle
LVH: Left ventricular hypertrophy
RWT: Relative wall thickness
TTE: Transthoracic echocardiography

## Acknowledgements

The authors thank the participants and staff of HCHS/SOL for their important contributions to this research. The views expressed in this manuscript are those of the authors and do not necessarily represent the views of the National Heart, Lung, and Blood Institute; the National Institutes of Health; or the U.S. Department of Health and Human Services.

## Pathophysiological Novelty and Relevance

### What is new?

- First study to evaluate the cardiac sequelae of de novo hypertensive disorder of pregnancy (HDP) in a cohort of 5,168 Hispanic/Latina women.

### What is relevant?

- Women with de novo HDP had detectable and measurable subclinical alterations in cardiac structure and both systolic and diastolic dysfunction, above and beyond the effects of current hypertension.

### Clinical/Pathophysiologic Implications?

- Our findings suggest that women with de novo HDP have pathophysiologic cardiac sequelae decades later that likely play a role in modulating long-term cardiovascular risk in women.

---

**Supplemental Table 1.** Differences in Measures of Left Ventricle Structure and Function by Type of Hypertensive Disorders of Pregnancy.

| Echocardiographic Measures | De Novo Hypertensive Disorders of Pregnancy History |  |  |  | p-trend |
| --- | --- | --- | --- | --- | --- |
|  | None | Gestational Hypertension | Preeclampsia | Eclampsia |  |
| **Continuous Measures** |  |  |  |  |  |
| LV ejection fraction, % | 71.72 (4.44) | 70.65 (4.65) | 71.22 (4.26) | 69.36 (6.23) | <0.001 |
| LV stroke volume, mL | 59.86 (11.15) | 63.64 (13.03) | 61.90 (11.56) | 59.34 (10.67) | <0.001 |
| LV mass index, g/m<sup>2</sup> | 75.61 (15.91) | 80.06 (20.75) | 75.70 (16.42) | 76.86 (18.35) | 0.19 |
| LV end-diastolic diameter, cm | 4.23 (0.36) | 4.28 (0.44) | 4.26 (0.35) | 4.19 (0.38) | 0.60 |
| LVMI/EDV ratio | 0.93 (0.23) | 0.91 (0.27) | 0.89 (0.21) | 0.91 (0.20) | 0.003 |
| LV relative wall thickness, cm | 0.44 (0.07) | 0.46 (0.07) | 0.44 (0.07) | 0.47 (0.09) | <0.001 |
| PTRV, cm/s | 219.72 (30.37) | 226.58 (29.95) | 221.93 (23.70) | 222.72 (27.09) | 0.12 |
| Lateral E/e' ratio | 8.73 (3.38) | 9.22 (3.85) | 8.41 (2.62) | 8.59 (2.73) | 0.01 |
| **Categorical Measures** |  |  |  |  |  |
| Concentric LV remodeling, % | 45.2 | 50.4 | 39.6 | 66.6 | 0.07 |
| Concentric LVH, % | 8.3 | 16.8 | 9.7 | 4.1 | 0.005 |
| Eccentric LVH, % | 2.3 | 5.1 | 0.6 | 4.2 | 0.99 |
| Abnormal LV geometry, % | 55.9 | 72.2 | 50.2 | 74.9 | <0.001 |
| Abnormal LV diastolic function, % | 35.3 | 38.7 | 29.7 | 32.7 | 0.38 |

**Supplemental Table 2.** Associations between Gestational Hypertension and Preeclampsia and Measures of Left Ventricle Structure and Function.

| Echocardiographic Measures | Gestational Hypertension |  |  |  |  |  |
| --- | --- | --- | --- | --- | --- | --- |
|  | Model 1 |  | Model 2 |  | Model 3 |  |
|  | Coefficient (95% CI) | P value | Coefficient (95% CI) | P value | Coefficient (95% CI) | P value |
| **Continuous Measures** |  |  |  |  |  |  |
| LV ejection fraction, % | -0.92 (-1.60, -0.24) | 0.008 | -0.77 (-1.45, -0.09) | 0.027 | -0.61 (-1.32, 0.09) | 0.09 |
| LV stroke volume, mL | 2.82 (0.95, 4.69) | 0.003 | 1.88 (0.11, 3.65) | 0.038 | 1.14 (-0.53, 2.81) | 0.18 |
| LV mass index, g/m<sup>2</sup> | 5.20 (2.00, 8.41) | 0.002 | 2.63 (-0.45, 5.71) | 0.09 | 1.76 (-1.38, 4.90) | 0.27 |
| LV end-diastolic diameter, cm | 0.17 (-0.57, 0.90) | 0.66 | 0.002 (-0.71, 0.72) | 1.0 | -0.27 (-0.96, 0.43) | 0.45 |
| LVMI/EDV ratio | 0.009 (-0.04, 0.06) | 0.69 | -0.005 (-0.05, 0.04) | 0.83 | -0.0006 (-0.05, 0.05) | 0.98 |
| LV relative wall thickness, cm | 0.27 (0.17, 0.38) | <0.001 | 0.18 (0.07, 0.29) | 0.002 | 0.12 (0.01, 0.23) | 0.03 |
| PTRV, cm/s | 9.07 (2.79, 15.35) | 0.005 | 4.82 (-1.64, 11.29) | 0.14 | 1.96 (-4.33, 8.26) | 0.54 |
| **Categorical Measures** |  |  |  |  |  |  |
| Concentric LV remodeling, % | 1.38 (0.97, 1.96) | 0.07 | 1.32 (0.92, 1.90) | 0.13 | 1.23 (0.85, 1.79) | 0.27 |
| Concentric LVH, % | 2.48 (1.60, 3.87) | <0.001 | 1.76 (1.09, 2.85) | 0.021 | 1.54 (0.92, 2.58) | 0.10 |
| Eccentric LVH, % | 2.24 (1.01, 4.98) | 0.048 | 1.80 (0.78, 4.18) | 0.17 | 1.89 (0.74, 4.79) | 0.18 |
| Abnormal LV geometry, % | 2.44 (1.69, 3.52) | <0.001 | 1.96 (1.34, 2.86) | <0.001 | 1.79 (1.19, 2.69) | 0.005 |
| Abnormal LV diastolic function, % | 1.86 (1.30, 2.66) | <0.001 | 1.34 (0.92, 1.94) | 0.12 | 1.17 (0.80, 1.71) | 0.42 |
| Echocardiographic Measures | Preeclampsia |  |  |  |  |  |
|  | Model 1 |  | Model 2 |  | Model 3 |  |
|  | Coefficient (95% CI) | P value | Coefficient (95% CI) | P value | Coefficient (95% CI) | P value |
| **Continuous Measures** |  |  |  |  |  |  |
| LV ejection fraction, % | -0.55 (-1.35, 0.24) | 0.17 | -0.46 (-1.25, 0.32) | 0.25 | -0.43 (-1.19, 0.33) | 0.27 |
| LV stroke volume, mL | 1.11 (-1.01, 3.22) | 0.31 | 0.54 (-1.50, 2.59) | 0.60 | 0.27 (-1.81, 2.34) | 0.80 |
| LV mass index, g/m<sup>2</sup> | 1.33 (-2.18, 4.84) | 0.46 | -0.34 (-3.37, 2.69) | 0.83 | -0.22 (-3.18, 2.74) | 0.88 |
| LV end-diastolic diameter, cm | -0.03 (-0.78, 0.72) | 0.94 | -0.11 (-0.84, 0.63) | 0.78 | -0.15 (-0.82, 0.51) | 0.65 |
| LVMl/EDV ratio | -0.01 (-0.05, 0.03) | 0.56 | -0.02 (-0.06, 0.02) | 0.27 | -0.02 (-0.06, 0.02) | 0.37 |
| LV relative wall thickness, cm | 0.08 (-0.06, 0.23) | 0.24 | 0.02 (-0.11, 0.15) | 0.75 | 0.008 (-0.12, 0.14) | 0.90 |
| PTRV, cm/s | 4.40 (-2.73, 11.53) | 0.23 | 1.41 (-5.97, 8.80) | 0.71 | 1.19 (-6.37, 8.75) | 0.76 |
| **Categorical Measures** |  |  |  |  |  |  |
| Concentric LV remodeling, % | 0.95 (0.58, 1.57) | 0.85 | 0.92 (0.56, 1.53) | 0.75 | 0.86 (0.53, 1.41) | 0.56 |
| Concentric LVH, % | 1.46 (0.73, 2.93) | 0.28 | 1.14 (0.57, 2.27) | 0.71 | 1.15 (0.57, 2.29) | 0.70 |
| Eccentric LVH, % | 0.29 (0.06, 1.40) | 0.12 | 0.26 (0.05, 1.28) | 0.10 | 0.27 (0.06, 1.33) | 0.11 |
| Abnormal LV geometry, % | 1.02 (0.62, 1.68) | 0.94 | 0.87 (0.53, 1.43) | 0.57 | 0.83 (0.50, 1.37) | 0.46 |
| Abnormal LV diastolic function, % | 1.08 (0.56, 2.07) | 0.83 | 0.88 (0.44, 1.79) | 0.73 | 0.78 (0.37, 1.64) | 0.51 |

